# Detection of hepatitis B virus-host junction sequences in urine of infected patients

**DOI:** 10.1101/2021.04.28.21254562

**Authors:** Selena Y. Lin, Yih-Ping Su, Evan R. Trauger, Benjamin P. Song, Emilie G. C. Thompson, Malcolm C. Hoffman, Ting-Tsung Chang, Yih-Jyh Lin, Yu-Lan Kao, Yixiao Cui, Hie-Won Hann, Grace Park, Wei Song, Ying-Hsiu Su

## Abstract

**Background & Aims:** Integrated hepatitis B virus (HBV) DNA, found in >85% of HBV-associated hepatocellular carcinomas (HBV-HCC), can play a significant role in HBV-related liver disease progression. HBV-host junction sequences (HBV-JS’s), created through integration events, have been used to determine HBV-HCC clonality. Here, we investigate the feasibility of analyzing HBV integration in a noninvasive urine liquid biopsy.

**Approach & Results:** Utilizing an HBV-targeted NGS assay, we first identified HBV-JS’s in 8 HBV-HCC tissues and designed short-amplicon junction-specific PCR assays to detect HBV-JSs in matched urine. We detected and validated tissue-derived junctions in 5 of 8 matched urine samples. Next, we screened 32 urine samples collected from 25 HBV-infected patients (5 with hepatitis, 10 with cirrhosis, 4 with HCC, and 6 post-HCC). Encouragingly, all 32 urine samples contained HBV-JS’s detectable by HBV-targeted NGS. Of the 712 total HBV-JS’s detected in urine, 351 were in gene-coding regions, 11 of which, including *TERT*, had previously been reported as recurrent integration sites in HCC tissue and were found in urine of cirrhosis or HCC patients only. The integration breakpoints of HBV DNA detected in urine were found predominantly (∼70%) at a previously identified integration hotspot, HBV DR1-2.

**Conclusions:** HBV viral-host junction DNA can be detected in urine of HBV-infected patients. This study is the first study to demonstrate the potential for a noninvasive urine liquid biopsy of integrated HBV DNA to monitor HBV-infected patients for HBV-associated liver diseases and the efficacy of antiviral therapy.

## INTRODUCTION

Hepatitis B virus (HBV) integration can occur early in the course of infection (1-3). Integration events are known to cause insertional mutagenesis, chromosomal instability, activation of oncogenes, and inflammation, and thus can play a critical role in liver disease progression and the development of hepatocellular carcinoma (HCC) (4). Integrated viral DNA has been detected in more than 85% of HBV-associated HCC (HBV-HCC) cases (5-8). Although HBV integration sites have been observed throughout the human genome, integration into known HCC driver genes (such as *TERT, MLL4* and *CCNE1*) has been reported repeatedly in patients with HCC (9).

One outcome of HBV DNA integration is the creation of a unique HBV-host junction sequence (HBV-JS) representing a specific molecular signature of the infected cell. HBV DNA integrations have been used to determine clonality of recurrent HCC tumors in tissues (10, 11). Recent studies have shown that integrated HBV DNA can be detected in the circulation (12-15). We have previously demonstrated that urine of HCC patients contains cell-free DNA (cfDNA) with HCC-associated modifications (16-19), whereas urine of HBV-infected patients contains fragmented (but not intact) HBV DNA (20). We hypothesize that urine can serve as a noninvasive liquid biopsy for the investigation of HBV integration in HBV-infected patients.

In this proof-of-concept study, we assessed the feasibility of detecting HBV-JS’s in urine from HBV-infected patients. We first identified HBV-JS’s in HCC tissue followed by detection in matched urine by sensitive junction-specific PCR assays. Next, we detected HBV-JS’s in urine from HBV-infected patients with hepatitis, cirrhosis, and HCC, including those undergoing recurrence monitoring (post-HCC), using an HBV-targeted NGS assay. These findings support the potential of urine as a noninvasive liquid biopsy for integrated HBV DNA, which can be used to monitor the dynamics of HBV integration in the infected liver and assess HBV treatment efficacy and disease progression.

## PATIENTS AND METHODS

### Study subjects and specimens

Two studies, A and B, were performed. Study A specimens included archived DNA isolated from eight pairs of matched HBV-HCC formalin-fixed paraffin-embedded tumor tissue and urine samples. The specimens were obtained from patients in stages I-II undergoing surgery (**Table 1)** at the National Cheng-Kung University Medical Center, Taiwan, as described previously (17, 19). Study B was performed on urine samples collected from 25 HBV-infected patients, including 5 with hepatitis, 10 with cirrhosis, 4 with HCC, and 6 post-HCC, at the Thomas Jefferson University Hospital (**Table 2**). Five patients (B7, B15, B17, B19, and B20) provided more than one urine collection over the course of the study, resulting in a total of 32 urine samples. All patient samples were collected with written informed consent and in accordance with the guidelines of the Institutional Review Board.

**Table 1.** Characteristics of archived DNA from matched HBV-HCC tissue and urine for study A.

| **Patient ID** | **Sex (M/F)** | **Cirrhosis** | **Tumor stage\*** | **Tumor size (cm)** |
| --- | --- | --- | --- | --- |
| A1 | M | - | 1 | 4.0 |
| A2 | M | + | 2 | 7.0 |
| A3 | M | + | 1 | 2.0 |
| A4 | M | + | 2 | 3.4 |
| A5 | M | - | 2 | 2.3 |
| A6 | F | - | 1 | 3.0 |
| A7 | M | - | 1 | 4.5 |
| A8 | F | - | 2 | 10.0 |

\* HCC tumors were staged using the tumor-node-metastasis (TNM) staging system. Patients ranged from 29-75 years of age.

**Table 2.** Clinical characteristics of patients in study B.

| DZ | Patient ID | Sex (M/F) | ALT (IU/L) | Serum AFP (ng/mL) | HBV serum viral load (IU/mL) | HBe Ag | HBe Ab | Tumor stage<sup>#</sup> | Tumor size (cm) |
| --- | --- | --- | --- | --- | --- | --- | --- | --- | --- |
| Hepatitis (n=5) | B1 | M | 8 | 1.2 | < 20 | - | - | NA |  |
|  | B2 | M | 58 | 3.3 | 105 | - | + |  |  |
|  | B3 | M | 24 | 1.5 | < 20 | - | + |  |  |
|  | B4 | M | 27 | 1.0 | < 20 | - | + |  |  |
|  | B5 | M | 77 | 3.9 | < 20 | - | + |  |  |
| Cirrhosis (n=10) | B6 | M | 40 | 2.8 | < 20 | - | + | NA |  |
|  | B7* | M | 22 | 2.8 | < 20 | - | - |  |  |
|  | B8 | M | 30 | 1.4 | < 20 | - | - |  |  |
|  | B9 | M | 31 | 1.8 | < 20 | - | + |  |  |
|  | B10 | M | 18 | 1.2 | < 20 | - | - |  |  |
|  | B11 | M | 22 | 1.8 | < 20 | - | + |  |  |
|  | B12 | M | 16 | 3.1 | < 20 | - | + |  |  |
|  | B13 | M | 16 | 1.1 | < 20 | - | - |  |  |
|  | B14 | M | 50 | 256.7 | 90 | + | - |  |  |
|  | B15* | M | 83 | 1.8 | < 20 | + | - |  |  |
| HCC (n=4) | B16 | M | 29 | 88.9 | < 20 | - | + | I | 1.4 |
|  | B17* | F | NA | 1.2 | < 20 | - | + | I | 0.7 |
|  | B18^ | M | 38 | 3045.0 | 50 | + | - | I | 4.8 |
|  | B19* | M | 23 | 756.4 | < 20 | - | + | I | 3.0 |
| Post-HCC (n=6) | B20* | M | 28 | 3.5 | < 20 | + | - | NA |  |
|  | B21 | M | 46 | 1.2 | < 20 | - | - |  |  |
|  | B22 | M | 28 | 2.5 | < 20 | - | - |  |  |
|  | B23 | M | 28 | 35.3 | NA | - | + |  |  |
|  | B24 | M | NA | 47.1 | < 20 | - | + |  |  |
|  | B25 | M | 50 | 5.5 | < 20 | - | + |  |  |

<sup>#</sup> HCC tumors were staged using the tumor-node-metastasis (TNM) staging system; \*more than one urine collection taken in the course of the study; ^, recurrence detected; NA, not available. Patients ranged from 51-78 years of age.

Additionally, eight HBV-negative normal urine donors ranging from 23 to 60 years of age from four male and four females served as a normal urine donor cohort.

### Urine collection and DNA isolation

Urine samples were collected and total urine DNA isolated as previously described (21). For study A, cfDNA was obtained by isolating the <1kb fraction from total urine DNA using carboxylated magnetic beads, a method previously developed by our laboratory (22). For study B, the JBS urine cfDNA isolation kit (JBS Science Inc, PA) was used according to the manufacturer’s specification.

### HBV-JS detection by HBV-targeted NGS assay and validation by PCR-Sanger sequencing

Tissue DNA was fragmented by sonication and subjected to Illumina NGS library preparation as previously described (23). Ten cycles of library amplification with Herculase II Fusion polymerase were performed (Agilent Technologies, Santa Clara, CA). Urine cfDNA samples underwent NGS library preparation using the ThruPLEX Tag-Seq kit (Cat# R400585, Takara Bio, Mountain View, CA), which is designed for adaptor ligation to double-stranded DNA and contains unique molecular tags (UMTs), according to manufacturer specifications. Library DNA was subjected to an HBV-targeted NGS assay following the manufacturer’s instructions (JBS Science Inc, PA). The captured library DNA was pooled for NGS analysis on the Illumina MiSeq platform (Penn State Hershey Genomics Sciences Facility at Penn State College of Medicine, Hershey, PA) or on the NovaSeq platform (Psomagen Inc, Rockville, MD).

The ChimericSeq analysis software we developed previously (24) was used to detect HBV-JS’s in NGS datasets. For NGS libraries containing UMTs, UMT family consolidation and consensus read generation were performed using the software package Connor (25). Briefly, UMT-containing reads were aligned to human and HBV reference genomes, and paired reads were grouped into families based on the mapping coordinates and combined UMT sequence. For each UMT family, a consensus read pair was generated. The consensus reads were then processed by ChimericSeq for HBV-JS identification.

To validate junctions identified by NGS, junction-specific primers were designed for the host and viral sequences for tissue and urine (**Supplemental Table 1**). Tissue primers generating amplicons larger than 100 bp for Sanger sequencing, while urine primers were designed to target the same junction sequence generating amplicons of 65 bp or less. The junction-specific PCR was performed with Hotstart Plus Taq Polymerase (Qiagen, Valencia, CA). PCR products were visualized by agarose gel electrophoresis on a 2.2% FlashGel DNA Cassette (Lonza Group, Basel, Switzerland) and subsequently subjected to Sanger sequencing, a nested PCR reaction, or restriction endonuclease (RE) digestion using enzymes obtained from New England Biolabs (Ipswich, MA), according to the manufacturer’s specifications.

## Data availability

The sequence data for the subjects studied in this work who consented to data archiving have been deposited in the European Genome–Phenome Archive (EGA), www.ebi.ac.uk/ega, which is hosted by the European Bioinformatics Institute (EBI) www.ebi.ac.uk (accession no. pending).

## Statistical analysis

The *Chi*-square test was performed for categorical variables. For all statistical tests, the level of significance was defined as *p* < 0.05 to determine whether differences were significant.

## RESULTS

### Detection of HBV integration in urine DNA by junction-specific PCR assays

To investigate if HBV-JS’s can be detected in urine, we identified eight pairs of archived DNA samples from matched tissue and urine from HBV-HCC patients (study A, **Table 1**), with each urine DNA isolate extracted from at least 1 mL of urine. We first detected HBV-JS’s in tissue DNA using an HBV-targeted NGS assay as described in Material and Methods. HBV integrations were detected in all eight HCC tumor DNA samples (**Table 3**). The integrated regions included the *TERT* gene, the most frequently reported integrated host gene (patient A5) (9). The junctions were validated by Sanger sequencing (**Supplemental Fig. 1**) using primers listed in **Supplemental Table 1**.

**Table 3.**
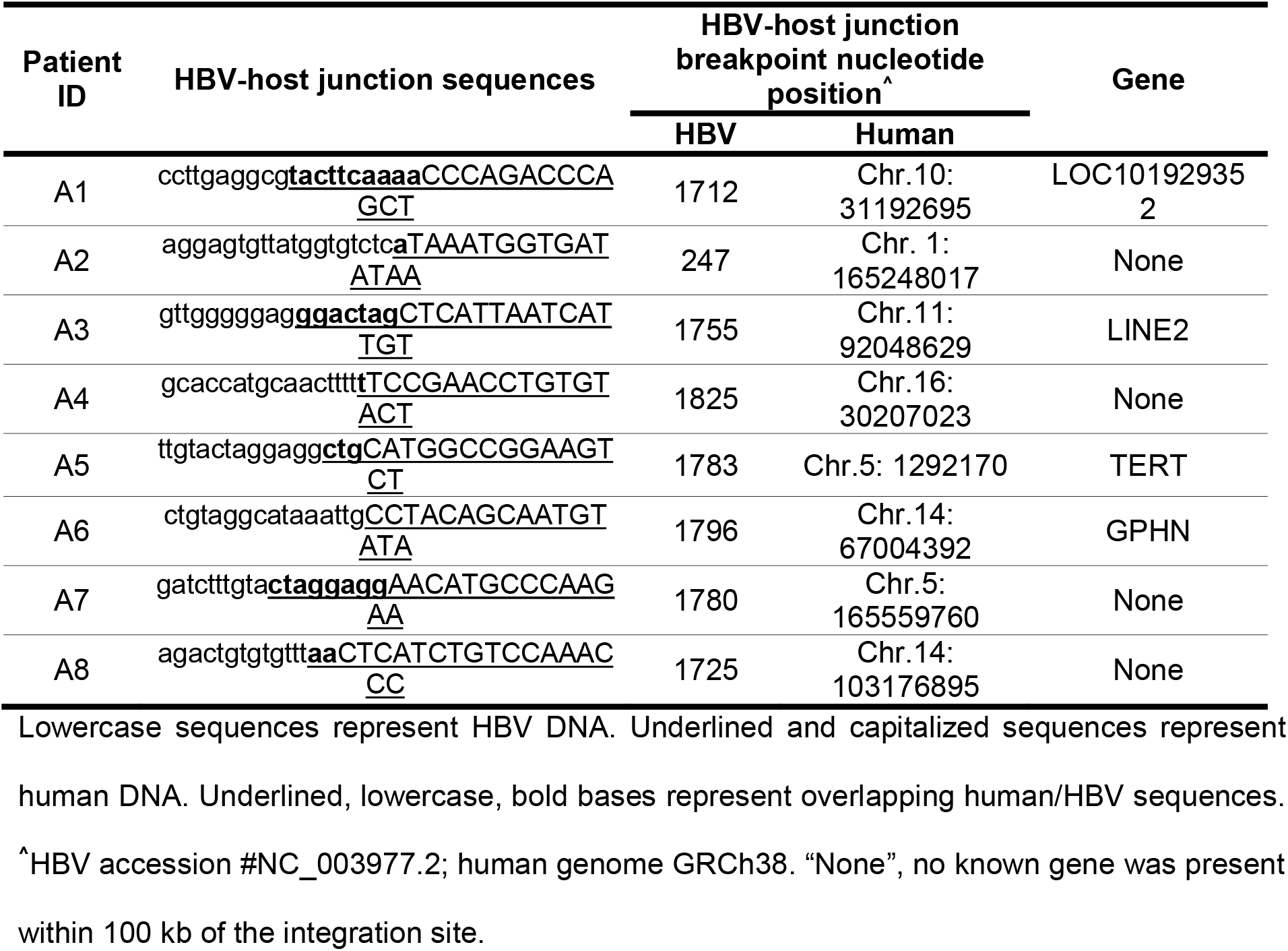
HBV integrations identified in archived tumor tissue DNA of study A patients.

To detect HCC tissue-derived HBV junction DNA in urine, we designed a junction-specific PCR assay for each junction (**Supplemental Table 1**). The amplicons were designed to be short since urine cfDNA is mostly fragmented to less than 250 bp (21). Matched tissue and urine DNA samples were subjected to junction-specific PCR amplification. PCR products of expected size were generated from five of the eight matched urine samples tested. To determine if the PCR products generated from the urine and tissue in each of the five tissue-urine pairs were identical, we used either two-step nested PCR followed by RE digestion of PCR products (patients A5, A6, and A8; **Fig. 1A**) or Sanger sequencing when no suitable RE site was available in PCR products (patients A1 and A2; **Fig. 1B-D**). Four (patients A2, A5, A6, and A8) of the five PCR product pairs contained sequences indistinguishable by the above methods, indicating that the HCC-tissue-derived HBV-JS was present in the matched urine.

**Figure 1.**
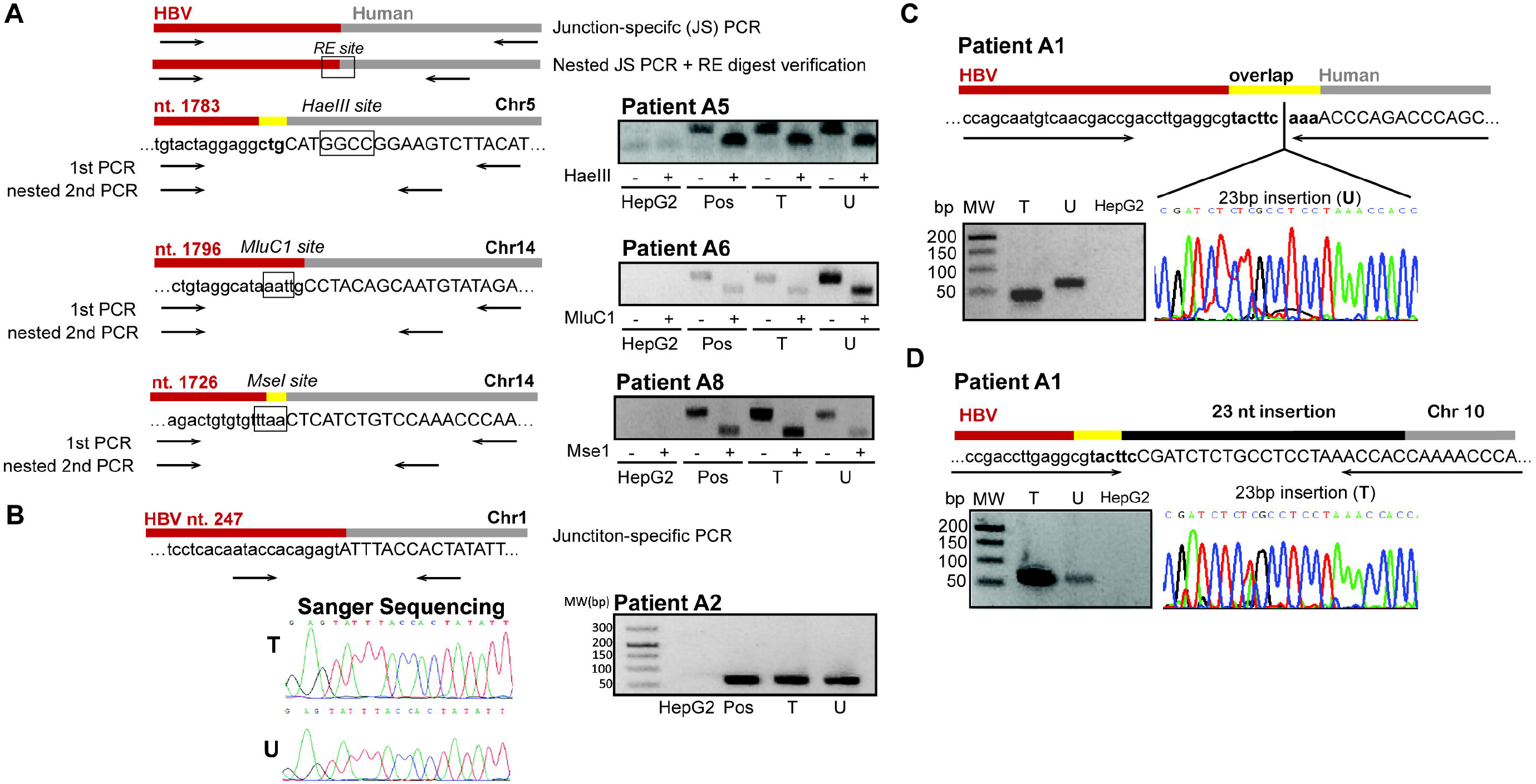
Detection of HBV-host junction sequences in study A, matched archived tissue and urine DNA. **(A)** Restriction endonuclease (RE) digestion approach. Tissue (T) and urine (U) DNA was amplified using two-step nested junction-specific PCR. The products generated by HBV and human primers were incubated in the absence (-) or presence (+) of the respective RE and analyzed by gel electrophoresis. The expected RE-digested PCR product sizes are patient A5: 25bp and 20bp of 45bp PCR product; A6: 24bp and 26bp of 53bp PCR product; A8: 22bp and 36bp of 57bp PCR product. **(B)** Sanger sequencing approach. The junction sequence of patient A2 was generated by PCR amplification of tissue and urine DNA using HBV and human primers. **(C)** Sequence of the HBV-host junction at chromosome 10 (Chr10) in urine of patient A1. Sanger sequencing of the urine-derived junction PCR product (U) revealed a 23-nt insertion compared with the tissue (T) product, as shown by gel electrophoresis. **(D)** Detection of the 23-nt inserted HBV-host junction in the corresponding tissue. Utilizing a hybrid Chr5-Chr10 primer, a specific tissue junction PCR product was obtained and found to be identical by sequencing to the urine-derived 23-nt insertion. Tissue DNA and HepG2 DNA (HepG2) DNA served as positive (Pos) and negative controls, respectively. MW denotes molecular weight marker.

Interestingly, the sizes of the junction PCR products from patient A1 were different between tissue and urine (**Fig. 1C**). Sanger sequencing analysis identified a 23-nucleotide (nt) insert indicating a re-arrangement between chromosomes 5 and 10 in the urine DNA sample. To determine if the urine-identified integration site could be found in tumor tissue DNA, we designed a primer across the chimeric chromosome 5/10 sequence (**Fig. 1D**) to specifically amplify the 23-nt-insert-containing HBV-JS species. Remarkably, the tripartite HBV-JS was successfully amplified and validated in the matched tissue DNA sample (**Fig. 1D)**. In total, we were able to detect and verify five of the eight HCC-tissue-derived HBV-JS’s in matched urine samples.

### Detection and characterization of HBV-JS’s in urine of HBV-infected patients by an HBV-targeted NGS assay

After detecting liver-derived HBV-JS’s in urine of HBV-HCC patients by junction-specific PCR assays, we next assessed if it was feasible to analyze HBV integrations in urine without prior knowledge of junction sequences from tumor tissue and before disease progression to HCC. A total of 32 urine specimens were collected from 25 HBV-infected individuals, including 5 patients with hepatitis, 10 with cirrhosis, 4 with HCC, and 6 post-HCC (study B; summarized in **Table 2)**. Five patients provided more than one urine specimen in the course of the study. Urine DNA was isolated from HBV-infected patients (Table 2) and normal donors, enriched for HBV sequences, and subjected to NGS followed by HBV-JS identification as described in Materials and Methods. Encouragingly, all 32 HBV-infected urine samples contained HBV-JS chimeric reads after UMT consolidation with at least 2 supporting reads (SR), a threshold applied for stringent HBV-JS identification and consistent with other reports (26-29).. Additionally, there were no detectable HBV-JS chimeric reads of ≥2 SR in the normal urine donor cohort.

Of all integration junctions identified in HBV-infected patient urine, a total of 712 HBV-JS’s from 31 urine samples These included 84, 366, 41, and 221 junctions detected in 4 hepatitis, 12 cirrhosis, 6 HCC, and 9 post-HCC urine samples, respectively. **Table 4** lists the major HBV-JS’s of each individual urine sample, defined as the two most abundant HBV-JS’s supported by at least 3% of total junction reads. As expected, we detected integrations in both gene-coding and non-coding regions in all disease categories. The latter included repeat regions, such as LINEs, SINEs, and simple repeats (30), that have been previously reported as HBV integration sites. Among the targets of the 351 unique integrations in gene-coding regions detected in urine, 11 genes had been previously reported in liver tissue from patients with severe liver diseases, such as cirrhosis and HCC. These genes are *AC007880*.*1, ADAM12, ARSA, ATF6, CLEC2L, FOCAD, PPP2R2C, TNFRSF9, TXNDC16, VEGFC*, and *TERT*. Most interestingly, all the genes, except for *AC007880*.*1* and *ARSA*, have been associated with the development of HCC (*ADAM12* (31), *ATF6* (32), *CLEC2L* (33) *TERT, VEGFC (34))* or other cancers (*FOCAD (35), PPP2R2C (36), TNFRSF9 (37), TXNDC16 (38))*.

**Table 4.** Major HBV integrations identified in urine samples of HBV-infected patients (study B).

| Patient ID | Diagnosis | # unique of HBV-JS | Major junctions (% of total junction reads)* |
| --- | --- | --- | --- |
| B1 | **Hepatitis (n=5)** | 48 | Chr4 (5.0%), Simple repeat (4.3%) |
| B2 |  | 4 | LINE (50.0%) |
| B3 |  | 2 | Chr1-a (50.0%), Chr1-b (50.0%) |
| B4 |  | 20 | CHCHD2P9 (20.7%), HSPE1P8 (17.2%) |
| B5 |  | 10 | SINE (19.0%), SEMA6D (14.3%) |
| B6 | **Cirrhosis (n=12)** | 21 | MLLT4 (10.2%), FOCAD (8.2%), PAK3 (8.2%) |
| B7a<sup>1</sup> |  | 21 | BAGE2 (6.8%), Chr12 (6.8%) |
| B7b |  | 5 | GCSHP2 (20.0%), PPP2R2C (20.0%) |
| B8 |  | 30 | Simple repeat (6.3%), LINE (4.8%) |
| B9 |  | 10 | Simple repeat (28.0%) |
| B10 |  | 40 | XRCC3 (9.6%), MST1L (8.0%) |
| B11 |  | 18 | TERT (48.8%), Chr16 (7.1%) |
| B12 |  | 53 | TERT (30.5%), GCSHP2 (6.9%) |
| B13 |  | 54 | Simple repeat (5.1%), LINC01481 (4.4%) |
| B14 |  | 13 | AL954742.1 (13.8%), Chr5 (10.3%) |
| B15a<sup>1</sup> |  | 46 | SINE (13.2%), LINE (3.9%) |
| B15b |  | 58 | SINE (8.0%), EXD3 (6.8%) |
| B16 | **HCC (n=6)** | 4 | KIF14 (25.0%), TET (25.0%) |
| B17a<sup>1</sup> |  | 2 | VPS8 (50.0%); Chr9 (50.0%) |
| B17b |  | 5 | Simple repeat (20.0%), AC011193.1 (20.0%) |
| B18 |  | 11 | Simple repeat (36.4%), TERT (9.1%) |
| B19a<sup>1</sup> |  | 17 | CTD-3229J4.1 (50.8%), Simple repeat (9.2%) |
| B19b |  | 2 | Simple repeat-a (50.0%); Simple repeat-b (50.0%) |
| B20a<sup>1</sup> | **Post-HCC (n=9)** | 16 | Simple repeat (25.0%), LTR (12.5%) |
| B20b |  | 63 | LMF1-AS1 (5.6%), hsd17b6 (5.1%) |
| B20c |  | 14 | GCSHP2 (60.8%), Simple repeat (10.1%) |
| B20d |  | 13 | Simple repeat (21.4%), LACTB2-AS1 (7.1%) |
| B21 |  | 4 | Simple repeat (75.0%) |
| B22 |  | 13 | TRPM3 (37.0%), ANKRD9 (13.0%) |
| B23 |  | 2 | PCDHGC5 (50.0%), HBB (50.0%) |
| B24 |  | 11 | Chr12 (20.7%), GALNT6 (10.3%) |
| B25 |  | 82 | LINE (12.4%), LTR (5.8%) |

\* Defined as up to two most abundant junctions with $\geq 2$ supporting reads; <sup>1</sup> multiple collections were obtained from the patient.

### Distribution of integrated HBV breakpoints detected in urine

We next examined the HBV genome breakpoints of the 712 junction sequences detected in urine. The HBV sequences of these junctions were plotted based on their locations in the HBV genome (**Fig. 2A**) and summarized in **Fig. 2B**. Approximately 70% of HBV DNA breakpoints detected in urine were clustered in the HBV DR1-2 region, a known integration hotspot (8) (**Fig. 2B**). The second most frequent of the previously reported HBV breakpoint regions, PreS (8), was also identified in urine, with higher frequencies found in hepatitis (7.1%), cirrhosis (10.5%), and post-HCC (7.3%) as compared with HCC (4.9%). However, the association of integrated HBV breakpoint location (DR1-2, PreS1/2, or other) with disease type was not statistically significant (χ^2^(6, 712)=6.3; *p*=0.39) (**Fig. 2B**).

**Figure 2.**
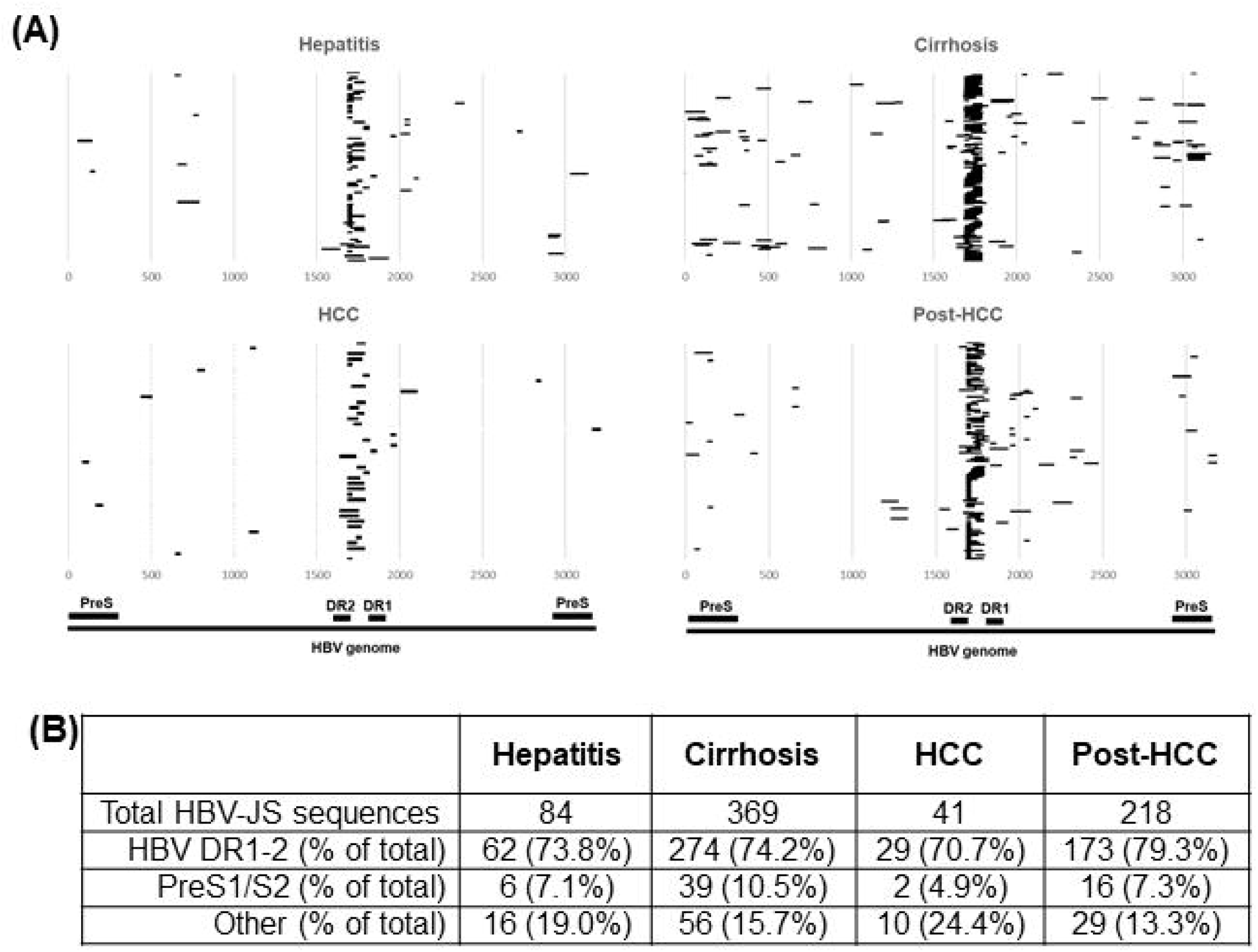
Analysis of HBV integrations in urine of HBV-infected patients with various liver diseases. **(A)** Distribution of HBV junction sequence breakpoints across the HBV genome. The HBV sequences indicated by black lines of unique junctions from each patient are plotted by disease category. Totals of 84, 366, 41, and 221 sequences from hepatitis, cirrhosis, HCC, and post-HCC samples, respectively, are shown. **(B)** Comparison of HBV DNA breakpoints in HBV DR1-2, PreS1/S2, and other HBV region among the HBV disease categories.

### Profiling of detectable HBV integrations sites in serially collected urine samples

Urine cfDNA containing HBS-JS’s may reflect the HBV integration profile of the infected liver. For a preliminary assessment of the possibility of using HBS-JS’s for disease monitoring, we collected serial urine samples from five patients (B7, B15, B17, B19, and B20) and compared the HBV-JS’s detected in different collections (**Table 4**). Patients B15 and B19, both with cirrhosis, had two collections one week apart and showed consistent integration in SINE and simple repeat regions, respectively, although they do not occur at the same integration breakpoint. Similarly, patient B20 (post-HCC) had four collections at 3-month intervals, all showing integration in non-coding regions (e.g., simple repeats), although the specific sites varied. Patient B17 (HCC) had two urine collections 2 months apart, with different junction sequences detected. Patient B7, with cirrhosis, had two urine samples collected 7 months apart, with integrations initially detected in a non-coding region followed by integrations in gene-coding regions. Interestingly, patient B7 developed HCC 4 years after the second collection. This initial comparison suggests that different HBV integrations can be captured in urine through serial monitoring. Collectively, our data demonstrate the feasibility of analyzing HBV integration using urine liquid biopsy.

## DISCUSSION

In this proof-of-concept study, we provide evidence for noninvasive detection of integrated HBV DNA in urine of HBV-infected patients with liver disease ranging from hepatitis to HCC. The novelty of this study is threefold. First, it is the first study to report detection of integrated HBV DNA in urine. Second, it includes the first comparative analysis of HBV integration in hepatitis, cirrhosis, HCC, and post-HCC using solely ectopic samples. Third, this study provides the first unambiguous evidence that urine contains liver-derived DNA, by taking advantage of the liver tropism of HBV and unique HBV-JS’s as molecular signatures of infected hepatocytes. Taken together, our results demonstrate the potential of urine liquid biopsy for integrated-HBV burden and HBV-related liver disease monitoring.

Our previous study demonstrated that only fragmented HBV DNA was detected in urine of patients with HBV infection even when the blood viral load was as high as 10^8^ copies/mL (20). Urine cfDNA is mostly mono- and dinucleosomal in size and largely derived from apoptotic cells (21). Therefore, HBV-sequence-containing urine cfDNA likely originates from degradation of host-integrated HBV genomes protected by nucleosomes in apoptotic HBV-infected hepatocytes. Hepatocytes from the HBV-infected liver have a higher turnover rate than hepatocytes from uninfected normal liver (39). Thus, urine provides a unique biopsy source to noninvasively detect integrated HBV DNA and monitor infected-cell burden, neither of which is achievable with current clinical HBV tests.

As expected, HBV genome integration breakpoints detected in urine were found to be mostly (>70%) in the HBV DR1-2 region, consistent with previous HBV-HCC tissue studies (8). Additionally, 11 HBV-JS’s identified in urine and reported previously in HCC tissue were all found in patients with cirrhosis or HCC. Urine DNA is likely to reflect the heterogeneity of HBV-JS’s in the liver more accurately compared with a single tumor biopsy or resection. HBV-JS’s have been detected in plasma of patients with HCC in recent studies (12-15). However, because urine cfDNA analysis is not confounded by the possible presence of replicating HBV or virions (20), urine is likely to provide a more sensitive liquid biopsy than blood for assessing integrated HBV DNA sequences in infected patients. Although we did not directly compare matched urine and plasma for HBV-JS detection due to plasma unavailability, the large number of HBV-JS’s we detected (712 in 32 urine samples) supports the notion that urine may be a more sensitive liquid biopsy for integrated HBV DNA.

Characterization of integrated HBV DNA and its urinary levels may provide insight into clonal expansion and turnover of infected hepatocytes, HCC risk stratification, monitoring recurrence post-treatment, and treatment efficacy. Frequent monitoring via urine liquid biopsy would be particularly advantageous in assessment of HBV-JS dynamics. The detection of HBV-host sequences and a chromosome rearrangement in HCC urine echoes our recent progress in developing a sensitive, noninvasive, urine-based HCC screening test (16, 19, 40). Further study with a much larger sample size is needed to determine the detection sensitivity and urine quantity needed for robust, reliable HBV-JS profiling and its clinical applications. In the present proof-of-concept study, we demonstrated that detection of HBV-JS’s in urine is feasible and holds potential for application in assessing HBV-related liver disease in infected patients.

## Data Availability

The sequence data for the subjects studied in this work who consented to data archiving have been deposited in the European Genome Phenome Archive (EGA), www.ebi.ac.uk/ega, which is hosted by the European Bioinformatics Institute (EBI) www.ebi.ac.uk (accession no. pending).

## List of Abbreviations

HBV: hepatitis B virus
HBV-HCC: HBV-infected hepatocellular carcinomas
HBV-JS’s: HBV-host junction sequences
HCC: hepatocellular carcinoma
cfDNA: cell-free DNA
UMT: unique molecular tag
RE: restriction endonuclease
nt: nucleotide

## References

1. Tu T, Budzinska MA, Vondran FW, Shackel NA, Urban SJJoV. Hepatitis B virus DNA integration occurs early in the viral life cycle in an in vitro infection model via NTCP-dependent uptake of enveloped virus particles. 2018.

2. Chauhan R, Churchill ND, Mulrooney-Cousins PM, Michalak TIJO. Initial sites of hepadnavirus integration into host genome in human hepatocytes and in the woodchuck model of hepatitis B-associated hepatocellular carcinoma. 2017;6:e317–e317.

3. Chauhan R, Shimizu Y, Watashi K, Wakita T, Fukasawa M, Michalak TIJCg. Retrotransposon elements among initial sites of hepatitis B virus integration into human genome in the HepG2-NTCP cell infection model. 2019;235:39–56.

4. Zhao K, Liu A, Xia YJTI. Insights into Hepatitis B Virus DNA Integration-55 years after virus discovery. 2020:100034.

5. Shafritz DA, Shouval D, Sherman HI, Hadziyannis SJ, Kew MC. Integration of hepatitis B virus DNA into the genome of liver cells in chronic liver disease and hepatocellular carcinoma. New England Journal of Medicine 1981;305:1067–1073.

6. Matsubara K, Tokino T. Integration of hepatitis B virus DNA and its implications for hepatocarcinogenesis. Molecular biology & medicine 1990;7:243.

7. Bonilla GR, Roberts LR. The role of hepatitis B virus integrations in the pathogenesis of human hepatocellular carcinoma. Journal of hepatology 2005;42:760.

8. Yan H, Yang Y, Zhang L, Tang G, Wang Y, Xue G, Zhou W, et al. Characterization of the genotype and integration patterns of hepatitis B virus in early and late onset hepatocellular carcinoma. 2015;61:1821–1831.

9. Li X, Zhang J, Yang Z, Kang J, Jiang S, Zhang T, Chen T, et al. The function of targeted host genes determines the oncogenicity of HBV integration in hepatocellular carcinoma. Journal of hepatology 2014;60:975–984.

10. Esumi M, Aritaka T, Arii M, Suzuki K, Tanikawa K, Mizuo H, Mima T, et al. Clonal origin of human hepatoma determined by integration of hepatitis B virus DNA. Cancer research 1986;46:5767–5771.

11. Esumi M, Tanaka Y, Tozuka S, Shikata T. Clonal state of human hepatocellular carcinoma and non-tumorous hepatocytes. Cancer chemotherapy and pharmacology 1989;23:S1–S3.

12. Li CL, Ho MC, Lin YY, Tzeng ST, Chen YJ, Pai HY, Wang YC, et al. Cell free virus host chimera DNA from Hepatitis B virus integration sites as a circulating biomarker of hepatocellular cancer. Hepatology 2020.

13. Chen W, Zhang K, Dong P, Fanning G, Tao C, Zhang H, Guo S, et al. Noninvasive chimeric DNA profiling identifies tumor-originated HBV integrants contributing to viral antigen expression in liver cancer. Hepatology International 2020:1–12.

14. Qu C, Wang Y, Wang P, Chen K, Wang M, Zeng H, Lu J, et al. Detection of early-stage hepatocellular carcinoma in asymptomatic HBsAg-seropositive individuals by liquid biopsy. Proceedings of the National Academy of Sciences 2019;116:6308–6312.

15. Li W, Cui X, Huo Q, Qi Y, Sun Y, Tan M, Kong Q. Profile of HBV integration in the plasma DNA of hepatocellular carcinoma patients. Current genomics 2019;20:61–68.

16. Hann H-W, Jain S, Park G, Steffen JD, Song W, Su Y-H. Detection of urine DNA markers for monitoring recurrent hepatocellular carcinoma. Hepatoma Research 2017;3:105–111.

17. Jain S, Xie L, Boldbaatar B, Lin SY, Hamilton JP, Meltzer SJ, Chen S-H, et al. Differential methylation of the promoter and first exon of the RASSF1A gene in hepatocarcinogenesis. Hepatology Research 2015.

18. Lin SY, Jain S, Song W, Hu C-T, Su Y-H. Strategic Assay Developments for Detection of HBV 1762T/1764A Double Mutation in Urine of Patients with HBV-Associated Hepatocellular Carcinomas. InTech Hepatocellular Carcinoma - Clinical Research 2012.

19. Lin SY, Dhillon V, Jain S, Chang T-T, Hu C-T, Lin Y-J, Chen S-H, et al. A locked nucleic acid clamp-mediated PCR assay for detection of a p53 codon 249 hotspot mutation in urine. The Journal of Molecular Diagnostics 2011;13:474–484.

20. Jain S, Su Y-H, Su Y-P, McCloud S, Xue R, Lee T-J, Lin S-C, et al. Characterization of the hepatitis B virus DNA detected in urine of chronic hepatitis B patients. 2018;18:40.

21. Su YH, Wang M, Brenner DE, Ng A, Melkonyan H, Umansky S, Syngal S, et al. Human urine contains small, 150 to 250 nucleotide-sized, soluble DNA derived from the circulation and may be useful in the detection of colorectal cancer. Journal of Molecular Diagnostics 2004;6:101–107.

22. Su Y-H, Song J, Wang Z, Wang X, Wang M, Brenner DE, Block TM. Removal of high molecular weight DNA by carboxylated magnetic beads enhances the detection of mutated K-ras DNA in urine. Annals of the New York Academy of Sciences 2008;1137:82–91.

23. Ding D, Lou X, Hua D, Yu W, Li L, Wang J, Gao F, et al. Recurrent Targeted Genes of Hepatitis B Virus in the Liver Cancer Genomes Identified by a Next-Generation Sequencing–Based Approach. PLoS Genetics 2012;8:e1003065.

24. Shieh F-S, Jongeneel P, Steffen JD, Lin S, Jain S, Song W, Su Y-H. ChimericSeq: An open-source, user-friendly interface for analyzing NGS data to identify and characterize viral-host chimeric sequences. PLOS ONE 2017;12:e0182843.

25. Michigan Uo. In: UM BRCF Bioinformatics Core.

26. Sung WK, Zheng H, Li S, Chen R, Liu X, Li Y, Lee NP, et al. Genome-wide survey of recurrent HBV integration in hepatocellular carcinoma. Nature genetics 2012;44:765–769.

27. Jiang Z, Jhunjhunwala S, Liu J, Haverty PM, Kennemer MI, Guan Y, Lee W, et al. The effects of hepatitis B virus integration into the genomes of hepatocellular carcinoma patients. Genome Research 2012.

28. Li W, Zeng X, Lee NP, Liu X, Chen S, Guo B, Yi S, et al. HIVID: an efficient method to detect HBV integration using low coverage sequencing. Genomics 2013;102:338–344.

29. Yan H, Yang Y, Zhang L, Tang G, Wang Y, Xue G, Zhou W, et al. Characterization of the genotype and integration patterns of hepatitis B virus in early and late onset hepatocellular carcinoma. Hepatology 2015;61:1821–1831.

30. Lee WY, Bachtiar M, Choo CC, Lee CGJBR. Comprehensive review of H epatitis BV irus associated hepatocellular carcinoma research through text mining and big data analytics. 2019;94:353–367.

31. Le Pabic H, Bonnier D, Wewer UM, Coutand A, Musso O, Baffet G, Clément B, et al. ADAM12 in human liver cancers: TGF β regulated expression in stellate cells is associated with matrix remodeling. 2003;37:1056–1066.

32. Shuda M, Kondoh N, Imazeki N, Tanaka K, Okada T, Mori K, Hada A, et al. Activation of the ATF6, XBP1 and grp78 genes in human hepatocellular carcinoma: a possible involvement of the ER stress pathway in hepatocarcinogenesis. 2003;38:605–614.

33. Zhao J, Ye J, Lin Y, Bu K, Mai R, Liu Z, Gao X, et al. Identification and validation of a ten-gene set variation score as a diagnostic and prognostic stratification tools in hepatocellular carcinoma. 2020.

34. Zhuang P-Y, Shen J, Zhu X-D, Lu L, Wang L, Tang Z-Y, Sun H-CJPo. Prognostic roles of cross-talk between peritumoral hepatocytes and stromal cells in hepatocellular carcinoma involving peritumoral VEGF-C, VEGFR-1 and VEGFR-3. 2013;8:e64598.

35. Weren RD, Venkatachalam R, Cazier JB, Farin HF, Kets CM, De Voer RM, Vreede L, et al. Germline deletions in the tumour suppressor gene FOCAD are associated with polyposis and colorectal cancer development. 2015;236:155–164.

36. Bluemn EG, Spencer ES, Mecham B, Gordon RR, Coleman I, Lewinshtein D, Mostaghel E, et al. PPP2R2C loss promotes castration-resistance and is associated with increased prostate cancer-specific mortality. 2013;11:568–578.

37. El Kramani N, Elsherbiny NM, El-Gayar AM, Ebrahim MA, Al-Gayyar MMJC. Clinical significance of the TNF-α receptors, TNFRSF2 and TNFRSF9, on cell migration molecules Fascin-1 and Versican in acute leukemia. 2018;111:523–529.

38. Wan F, Zhu Y, Han C, Xu Q, Wu J, Dai B, Zhang H, et al. Identification and validation of an eight gene expression signature for predicting high Fuhrman grade renal cell carcinoma. 2017;140:1199–1208.

39. Summers J, Jilbert AR, Yang W, Aldrich CE, Saputelli J, Litwin S, Toll E, et al. Hepatocyte turnover during resolution of a transient hepadnaviral infection. 2003;100:11652–11659.

40. Kim AK, Hamilton JP, Lin SY, Chang T-T, Hann H-W, Hu C-T, Lou Y, et al. Urine biomarker: novel approach to hepatocellular carcinoma screening. 2020:2020.2011.2021.20236125.

