## Supplemental File for "Detection of hepatitis B virus-host junction sequences in urine of infected patients"

### SUPPLEMENTARY FIGURES

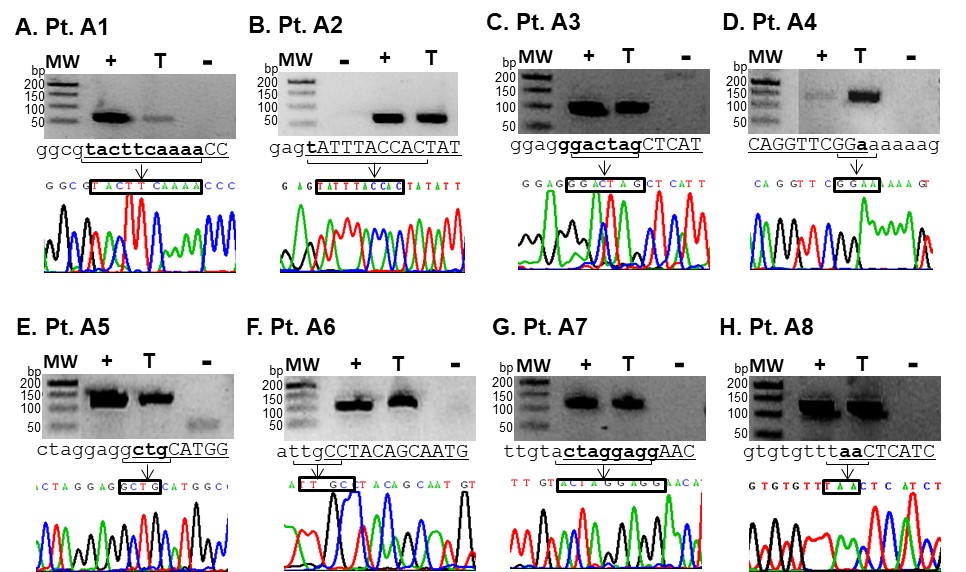

Supplemental Figure 1. Validation of NGS identified HBV-host junctions from archived HBV-HCC tissue DNA. Tissue DNA from patients was subjected to PCR amplification using primers for the major junction sequences identified from NGS analysis (gel image in each panel). The HBV-enriched tissue DNA library from the original NGS assay was used as a positive control (+) and HepG2 DNA as a negative control (-). Each PCR product was Sanger-sequenced, the junction sequence is denoted by the box in each electropherogram. Lower-case sequences represent HBV DNA. Underlined and capitalized sequences represent human DNA. Underlined, lower-case, bold sequences represent overlapping human/HBV sequences. MW, molecular weight ladder. Pt, patient.

### SUPPLEMENTARY TABLES

#### Supplemental Table 1. PCR primers used in the HCC matched tissue (T)-urine (U) PCR validation assays for study A.

| **Patient ID** | **Assay (T/U)** | **Sequence (5’ -3’)** |
| --- | --- | --- |
| A1 | PCR-Sanger sequencing (T) | GGGTCTGGGTTTTGAAGTA*CGCC* |
|  |  | GACCACCGTGAACGCCC |
|  | Short amplicon PCR (T/U) | CTGAGCTGGGTCTGGGTTTT |
|  |  | GTCTGGGTTTTGGTGGTTTAGGAG* |
|  |  | TGTCAACGACCGACCTTGAGGC |
| A2 | PCR-Sanger sequencing (T/U) | CCTGTGCTAGACCTGGGAATATAGTGGTAA |
|  |  | TGTTGACAAGAATCCTCACAATACCAC^#^ |
| A3 | PCR-Sanger sequencing (T) | GTGCCAGGTTTGACACAATG |
|  |  | AACGACCGACCTTGAGGCATACTTC |
|  | Short amplicon PCR (T/U) | GTGCCAGGTTTGACACAATGA |
|  |  | ACTGAGTGGGAGGAGTTGGG |
| A4 | PCR-Sanger sequencing (T) | CTTGGCCATTGGCTCTATGT |
|  |  | AATTGGTCTGTTCACCAGCA |
|  | Short amplicon PCR (T/U) | TTTAGTACACAGGTTCGGAAAAA |
|  |  | AATTGGTCTGTTCACCAGCA |
| A5 | PCR-Sanger sequencing (T) | CTCCCCACAAACTCCCAAG |
|  |  | AACGACCGACCTTGAGGCATACTTC |
|  | Short amplicon PCR (T/U) | CTCCCAAGACATGTAAGACTTCC |
|  |  | TGTACTAGGAGGCTGCATGG^#^ |
| A6 | PCR-Sanger sequencing (T) | CAGTAGGTCATAGTAAGTCAGCAAGC |
|  |  | TRGGGGAGGAGATAAGGTTAAAGGTC |
|  | Short amplicon PCR (T/U) | TAAAATCTATACATTGCTGTA*GGCA* |
|  |  | CTTTGTACTAGGAGGCTGTAGGCA |
| A7 | PCR-Sanger sequencing (T) | TTGGGCATGTTCCTCCTAG*T* |
|  |  | TGTCAACGACCGACCTTGAGGC |
|  | Short amplicon PCR (T/U) | TTGGGCATGTTCCTCCTAGT |
|  |  | GGGGAGGAGATTAGGTTAATGA |
| A8 | PCR-Sanger sequencing (T) | AACAAACCGTCACTTTCGTTG |
|  |  | CTCTTGGACTTTCGGCAATG |
|  | Short amplicon PCR (T/U) | TTTGGGTTTGGACAGATGAG |
|  |  | CCGACCTTGAGGCATACTTC |

* Primer sequence covers a rearranged human sequence; ^#^ contains human sequences in HBV primer; R is a degenerate base (A/G).
